# The “Double Eights Mask Brace” Improves the Fit and Protection and Protection of a Basic Surgical Mask Amidst Covid-19 Pandemic

**DOI:** 10.1101/2020.05.18.20099325

**Authors:** Daniel P. Runde, Karisa K. Harland, Paul Van Heukelom, Brett Faine, Patrick O’Shaughnessy, Nicholas M. Mohr

**Author notes:** **Corresponding Author Contact Information** Daniel P. Runde, University of Iowa Carver College of Medicine; 200 Hawkins Dr., 1008 RCP; Iowa City, IA, 52242.

## Abstract

**Study Objective:** The COVID-19 pandemic has resulted in widespread shortages in personal protective equipment, including N95 respirators. While basic surgical facemasks are more commonly available, their efficacy is limited due primarily to their poor face seal. This pilot study examined the impact of a rubber band mask brace on a basic surgical mask, as determined by quantitative fit testing.

**Methods:** Subjects wearing a basic surgical facemask and the rubber band mask brace underwent quantitative fit testing using machinery designed to certify N95 mask fit. Subjects were tested with the brace anchored behind their eyes, with a paperclip behind the head, and on the side knobs of their face shields. The primary outcome measure was whether the subject passed the quantitative fit test at or above the OSHA verified standard for N95 masks.

**Results:** Subjects (n=11) were 54.5% female, with a median height of 70 inches (IQR 68-74), weight of 170 lbs (IQR 145-215) and BMI of 24.6 (IQR 22.2-27.2), and encompassing 5 distinct N95 mask fit types. We found that 45%, 100% and 100% of subjects passed the quantitative fit test when the brace was anchored behind the ears, with a paperclip and on a face shield respectively.

**Conclusion:** Of the 11 subjects included in the analysis, across a range of body habitus and N95 mask fit types, all passed the quantitative fit test when the mask brace was anchored on either face shield or with a paperclip. This data suggests the brace would offer an improved margin of safety when worn with a basic surgical mask.

## Introduction

The COVID-19 pandemic has resulted in widespread shortages in personal protective equipment (PPE), including N95 respirators used by frontline healthcare workers.^1-3^ While basic surgical facemasks are more commonly available, their efficacy is limited due primarily to their poor seal, increasing exposure to aerosolized droplets, as these droplets will follow the path of least resistance, which is definitionally anywhere the mask does not have a good seal with the face, negating the filtering capabilities of the mask itself. We present an improvement which can be worn with a basic surgical mask, comprised of three interlocking rubber bands, that substantially improves the facial seal of the mask and may allow it to serve as a substitute for the N95 respirator in times of shortage. We refer to this improvement as the Double Eights Mask Brace (DEMB).

## Methods

Subjects wore an ASTM Level 1 basic surgical mask (Halyard 47567) accompanied by the DEMB. The brace consists of a central size #64 (3.5 x 0.25 inch) rubberband, which is fitted over the bridge of the nose and under the chin, and two size #33 (3.5 x 0.125 inch) rubber bands knotted together with the central band, which serve as anchors for the brace (see Fig. 1, model is author BF). The masks were fitted with a grommet and then subjected to OSHA standard quantitative fit testing (29 CFR 1910.134). Using the TSI PortaCount Pro Model 8038 and Model 8026 Particle Generator (Shoreview, MN), ambient saline particles were generated within the testing area and the concentration of these particles were sampled both inside and outside the mask (see Fig. 2). The external and internal measurements were continuously compared while the subject underwent the OSHA specified testing components: normal breathing, deep breathing, rotating the head side to side, flexing and extending the neck, talking while reading a pre-specified script, standing and bending continuously from the waist, and finally normal breathing again. The degree of mask seal present during each testing component was recorded, as was the overall fit factor of the mask. All evaluations were performed at the level of N95 respirator fit test standards, which requires an overall fit factor ≥ 100 on a 0-200 scale. For reference, the fit factor is expressed as the challenge aerosol concentration outside the respirator divided by the challenge aerosol concentration that leaks inside the respirator during the fit test. Subjects were tested in three scenarios: with the fit aid anchored behind the ears, behind the head with a paperclip, or on the side knobs of their face shields. In addition to whether the subject passed the quantitative fit test in each scenario, data on overall fit factor and the fit factor for the seven individual components of the test were recorded. Baseline characteristics were analyzed with summary statistics, and fit testing was compared between the 3 securement locations.

**Figure 1:**
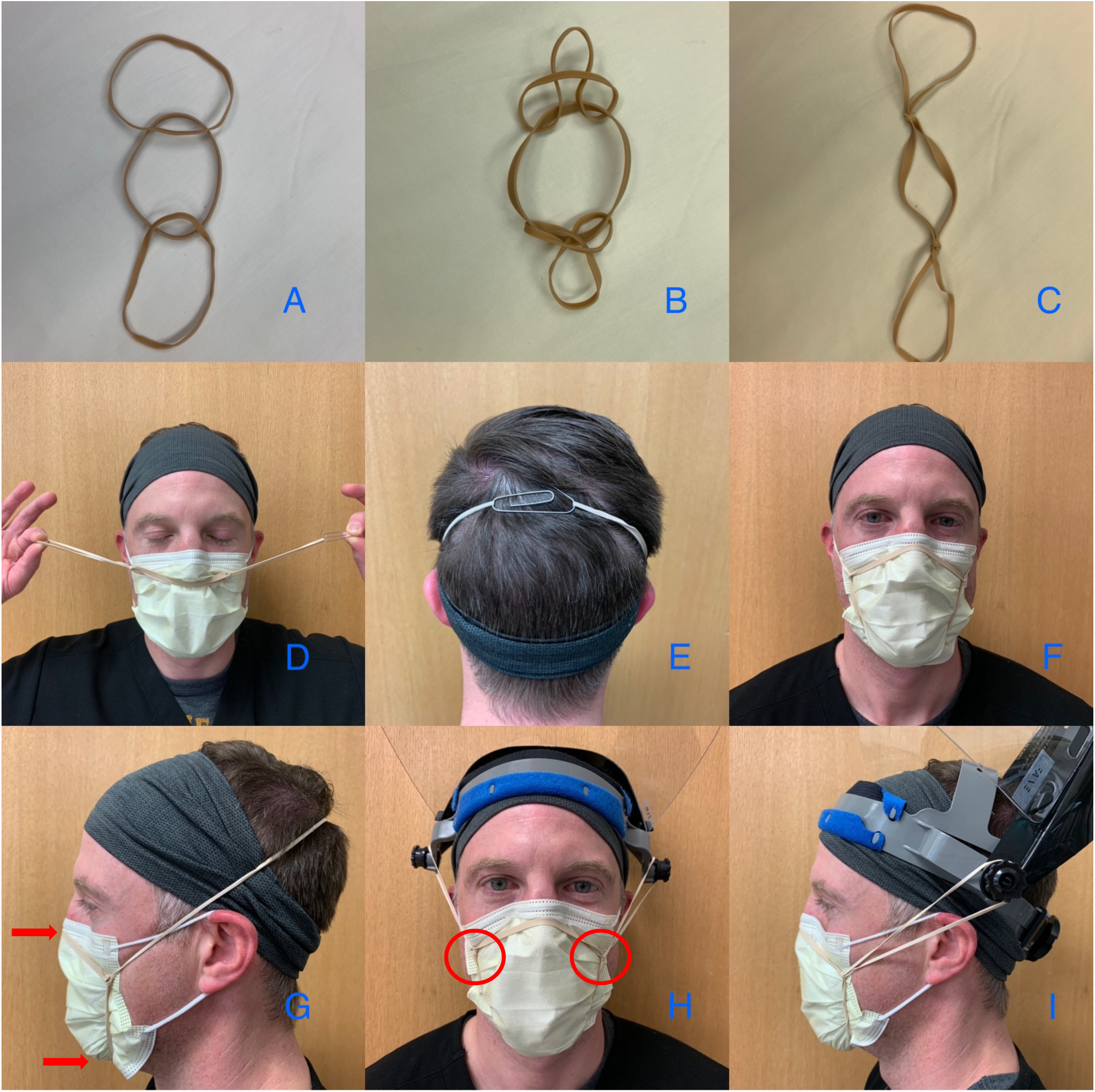
A - C demonstrate the way that the three rubber bands are knotted together. D - G demonstrate how to anchor and fit over a mask using a paper clip. H - I demonstrate how to fit and anchor the rubber bands using a face shield with side knobs. The red arrows in image G demonstrate proper positioning on the bridge of the nose and below the chin. The red circles in image H indicate proper position of the brace knots inside the edges of the mask.

**Figure 2:**
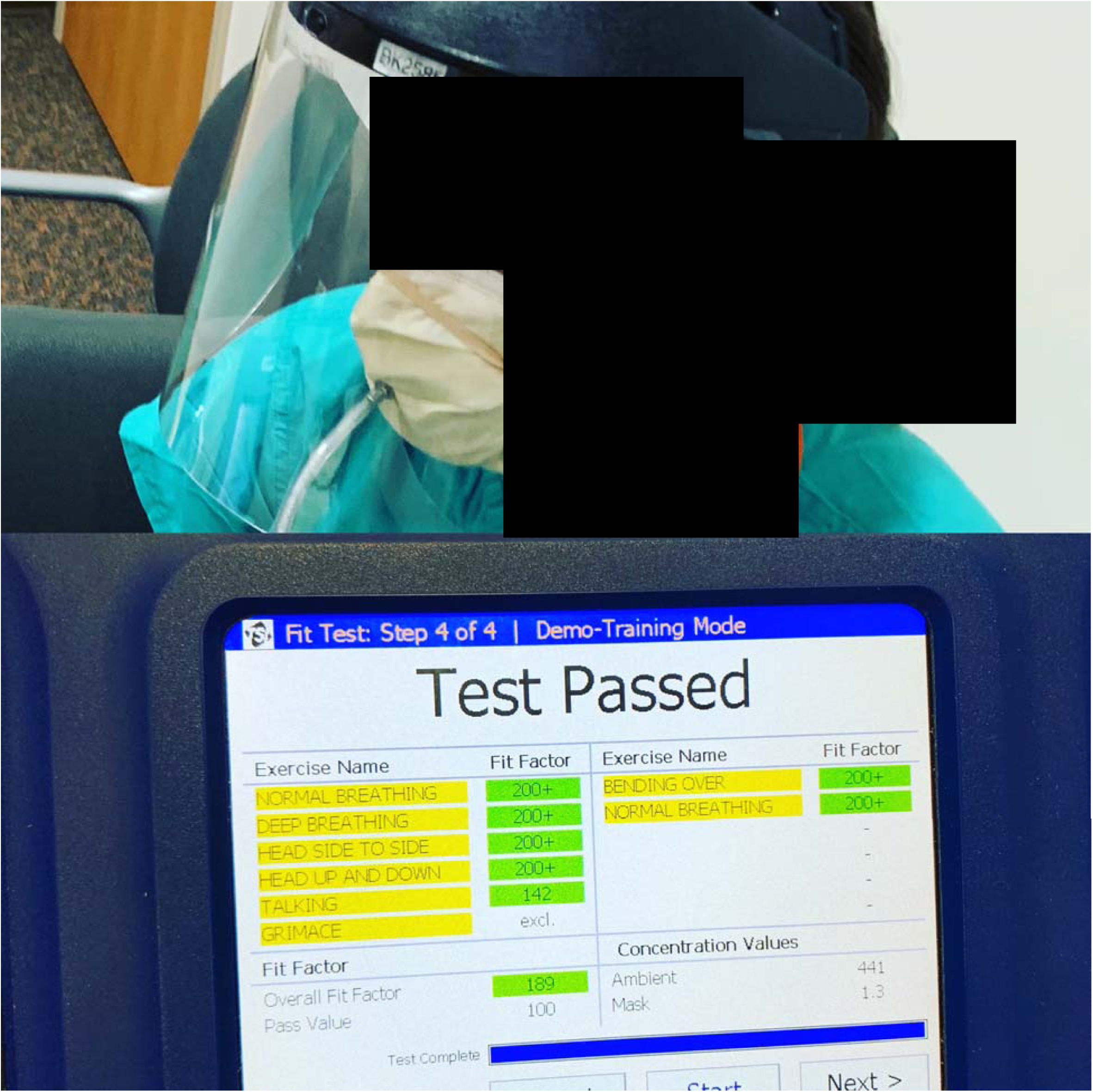
Halyard 47567 yellow mask fitted with a grommet and connected to the TSI PortaCount Pro along with results of quantitative fit testing

**Figure 3.**
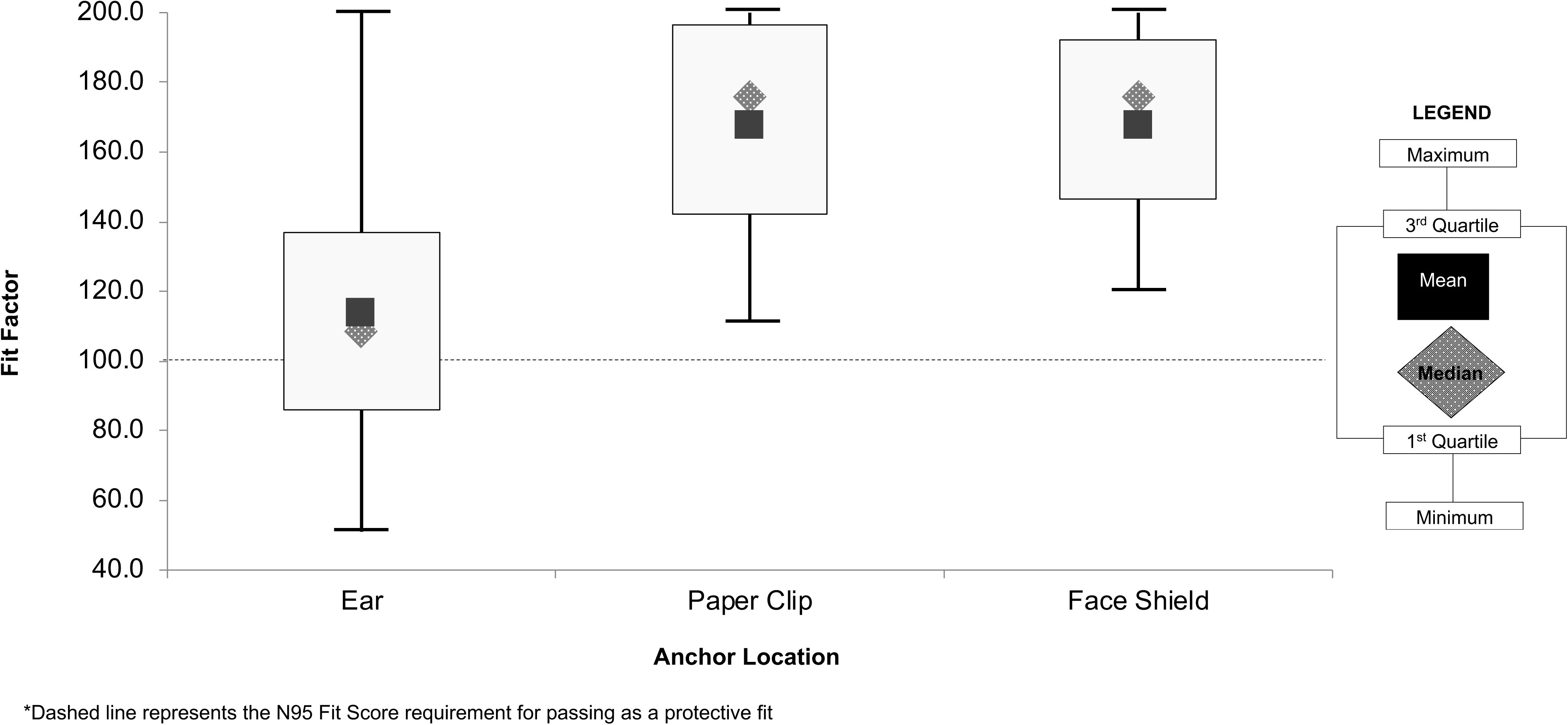
Distribution of Fit Factor by Mask Fit Anchor Type Among Healthcare Providers Fit for an N95.

### Filter Pressure Drop Testing

Filter pressure drop, or "breathing resistance,” measurements were conducted using a pass-through cylinder with an inner diameter of 2.7 cm. A portion of the surgical mask was tightly clamped between the upper and lower portions of the cylinder. The air flow rate through the cylinder was induced with a vacuum pump such that the velocity of air going through the mask portion was the same as if testing the entire mask at inflow rates of 15 - 85 LPM. These inflow rates represent the range of minute volumes experienced by a sedentary to a highly active person.^4^ Furthermore, 85 LPM is equivalent to the testing flow rate when performing the NIOSH certification test for N95s. The velocity calculation was conducted by comparing the area of the inner hole of the testing column with the measured area between the rubber bands encircling the nose and mouth of 154 cm2. Flexible tubing was used to connect a calibrated pressure transmitter (Series 646, Dwyer Instruments, Inc., Michigan City, MI) that measures in the range of 0 – 65 mm H2O air pressure to pressure taps installed above and below the mask media. NIOSH stipulates that an N95 should not exceed 30 mm H2O when tested at 85 L/min and this pressure level was used as the standard for the Halyard 47567 mask used in this study. The voltage output signal of the transmitter was received by an analog-to-digital converter and read using the Labview software system (National Instruments, Austin, TX) every 0.25 s. The average of 100 such measurements established the pressure drop across the mask section.

## Results

Subjects (n=11) were 54.5% female, with a median height of 70 inches (IQR 68-74), weight of 170 lbs (IQR 145-215) and BMI of 24.6 (IQR 22.2-27.2), and encompassing 5 distinct N95 mask fit types (1804S, 1860 Regular, 1870, 1870+ and Vflex 1805). Results demonstrated that 45%, 100% and 100% of subjects passed the quantitative fit test when the brace was anchored behind the ears, with a paperclip and on a face shield respectively, with mean/median overall fit factors of 114/108.5, 167.9/176 and 167.8/176 (Figure 2).

Pressure drop ranged between 1.3 – 7.7 mm H20 across the range of tested flow rates. These values demonstrate that breathing through the average 154 cm2 area circumscribed by the DEMB produces very reasonable breathing resistance even at high exertion breathing (85 LPM minute volume).

## Limitations

There are several limitations of this study. First, it is an exploratory study of a small number of subjects in a convenience sample. Findings of this single-site study may not be applicable to all types of basic surgical masks or to every type of facial morphology. It is also possible that heterogeneity in the specific available rubber bands could lead to variable brace performance. Finally, only one surgical mask type was evaluated quantitatively and the performance of the DEMB in a clinical setting was not evaluated.

## Discussion

Of the 11 subjects included in the analysis, across a range of body habitus and N95 mask fit types, all passed the quantitative fit test when the DEMB was anchored on the face shield or behind the head with a paperclip, suggesting that the implement would offer a substantially augmented degree of facial seal, to the point that it was able to meet the rigorous standards applied to fit tested N95 masks. Even the less robust performance of the mask brace when anchored behind the ears appears to offer improved seal compared to wearing the mask alone.

ASTM Level 1 masks (standard surgical masks) are certified to have a minimum bacterial filtration and sub-micron particulate filtration efficiency at 0.1 micron of ≥ 95%^5^. Importantly, even poorly performing filters have been shown to have near 100% efficiency for particles approaching 5 μm in diameter^6^, which covers the range of 5-10 μm respiratory droplets widely considered to be the primary route of SARS-CoV-2 transmission^7^.

Given the high burden of health care worker infection and the critical shortage in PPE, the dramatic improvement in mask face seal created by this affordable and practical mask brace offers an alternative protective device that has the potential to prevent transmission and save lives.

Daniel Runde, MD, MME

Karisa Harland, PhD

Paul Van Heukelom, MD

Brett Faine, PharmD, MS

Patrick O’Shaughnessy, PhD

Nicholas Mohr, MD, MS

## Data Availability

Data is available for review upon request.

## Acknowledgements

Michelle Duong Davenport, PhD and her collaborators at FixTheMask.com for creating the original concept for this mask brace. Barb Schuessler, MSN, RN, MBA, CPEN and Rachel Lewis, RN of the University of Iowa Hospitals and Clinics for providing access to and training on the quantitative fit testing equipment used in this study.

## Funding Sources/Disclosures

None

## Conflicts

The authors have no conflicts to report.

## Author contributions

DPR and PV designed the study. KKH, NM, BF and PO provided expertise in the interpretation of the data. BF posed as the model for Figure 1. DR, NM and KKH were responsible for managing the data and provided oversight of statistical analyses. DPR and KKH had full access to the data set and analyzed the data. DPR and KKH drafted the manuscript, and all authors contributed substantially to its revision. DPR takes responsibility for the paper as a whole.

## Notes

### Competing Interest Statement

The authors have declared no competing interest.

### Funding Statement

No external funding was received for this work.

